# Widespread Contamination of SARS-CoV-2 on Highly Touched Surfaces in Brazil During the Second Wave of the COVID-19 Pandemic

**DOI:** 10.1101/2021.06.14.21258894

**Authors:** Severino Jefferson Ribeiro da Silva, Jéssica Catarine Frutuoso do Nascimento, Wendell Palôma Maria dos Santos Reis, Caroline Targino Alves da Silva, Poliana Gomes da Silva, Renata Pessôa Germano Mendes, Allyson Andrade Mendonça, Bárbara Nazly Rodrigues Santos, Jurandy Júnior Ferraz de Magalhães, Alain Kohl, Lindomar Pena

**Affiliations:** Laboratory of Virology and Experimental Therapy (LAVITE), Department of Virology, Aggeu Magalhães Institute (IAM), Oswaldo Cruz Foundation (Fiocruz), 50670-420, Recife, Pernambuco, Brazil; Department of Virology, Pernambuco State Central Laboratory (LACEN/PE), Recife, Pernambuco, Brazil; University of Pernambuco (UPE), Serra Talhada Campus, Serra Talhada, Pernambuco, Brazil; MRC-University of Glasgow Centre for Virus Research, Glasgow, G61 1QH, UK

**Keywords:** SARS-CoV-2, Coronavirus disease 2019, Environmental contamination, Prevention policies, Transmission

## Abstract

Although SARS-CoV-2 surface contamination has been investigated in temperate climates, few studies have been conducted in the tropics. Here, we investigated the presence of SARS-CoV-2 on high-touch surfaces in a large city in Brazil. A total of 400 surface samples were collected in February 2021 in the City of Recife, Northeastern Brazil. A total of 97 samples (24.2%) tested positive for SARS-CoV-2 by RT-qPCR using the CDC-USA protocol. All the collection sites, except one (18/19, 94.7%) had at least one environmental surface sample contaminated. SARS-CoV-2 positivity was higher in public transport terminals (47/97, 48.4%), followed by health care units (26/97, 26.8%), public parks (14/97, 14.4%), public markets (4/97, 4.1%), and beach areas (4/97, 4.1%). Toilets, ATMs, handrails, playground, and outdoor gym were identified as fomites with the highest rates of viral contamination. Regarding the type of material, SARS-CoV-2 RNA was found more commonly on metal (45/97, 46.3%), followed by plastic (18/97, 18.5%), wood (12/97, 12.3%), rock (10/97, 10.3%), concrete (8/97, 8.2%), and glass (2/97, 2.0%). Taken together, our data indicated extensive SARS-CoV-2 contamination in public surfaces and identified critical control points that need to be targeted to break SARS-CoV-2 transmission chains.

**Synopsis:** We investigated the presence of SARS-CoV-2 on high-touch surfaces in a large city in Brazil and identified critical points to establish effective control measures aimed at breaking transmission.

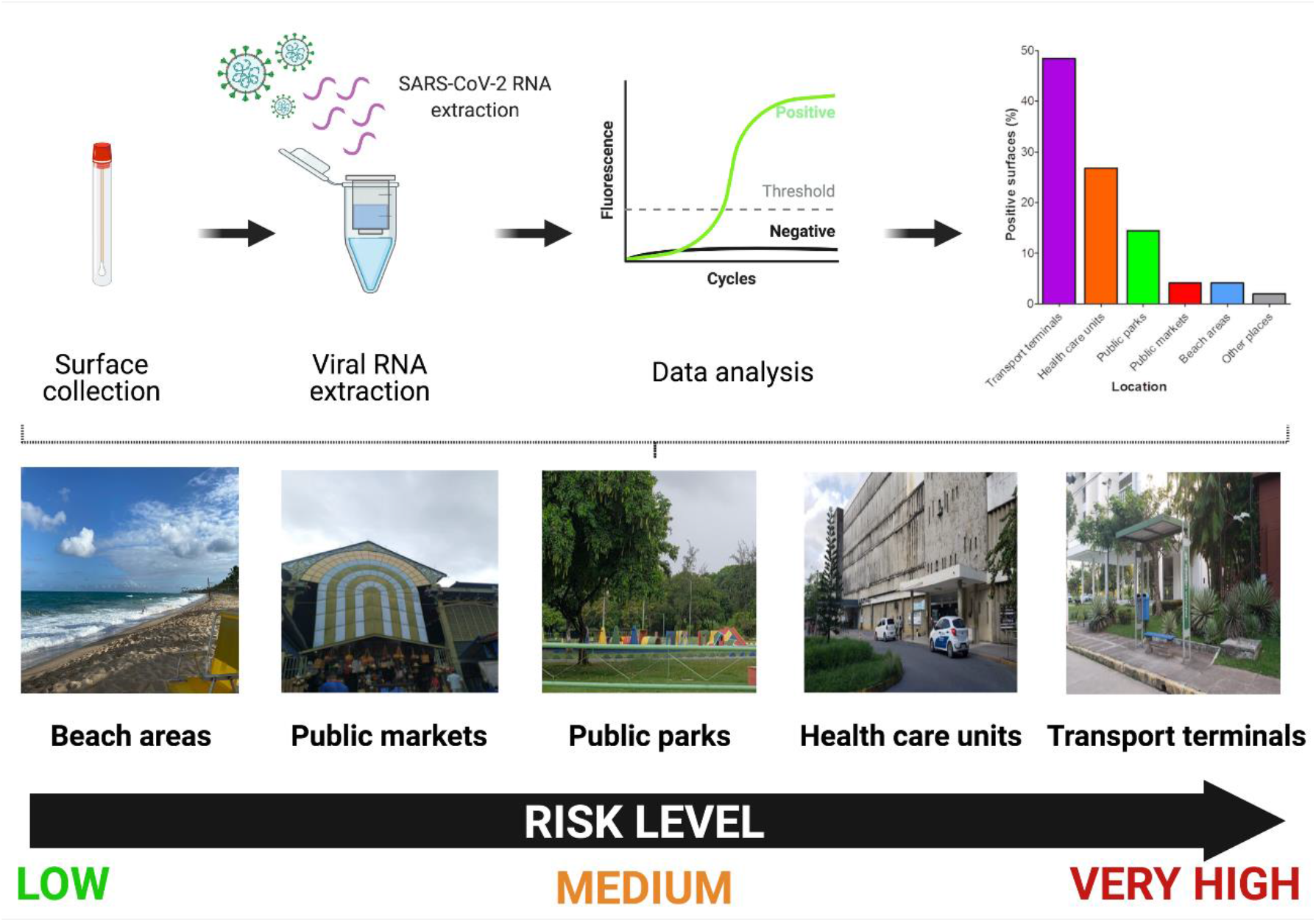

## INTRODUCTION

Coronaviruses (CoVs) are members of the *Coronaviridae* family and represent a diverse group of viruses that cause respiratory and intestinal infections in animals and humans ^1^. The *Coronavirinae* subfamily is divided into four genera - *Alphacoronavirus, Betacoronavirus, Gammacoronavirus, and Deltacoronavirus*. Alphacoronaviruses (HCoV-229E and HCoV-NL63) and Betacoronaviruses (HCoV-OC43 and HCoV-HKU1) are commonly associated with mild respiratory disease in humans ^2^. However, in the last two decades, three highly pathogenic betacoronaviruses have emerged from animal sources to cause severe respiratory disease in humans: severe acute respiratory syndrome coronavirus (SARS-CoV) ^3^, Middle East respiratory syndrome coronavirus (MERS-CoV) ^4^, and more recently, the severe acute respiratory syndrome coronavirus 2 (SARS-CoV-2)^5-7^.

SARS-CoV-2 first emerged in the city of Wuhan, Hubei province, China, in December 2019 causing an outbreak of a yet unknown acute pneumonia ^8^. Unlike SARS-CoV and MERS-CoV, the new coronavirus was found to be highly transmissible among humans and has spread rapidly around the globe prompting the World Health Organization (WHO) to declare a pandemic on March 11, 2020 (WHO, 2020). As of June 7, 2021, there have been approximately 173.4 million confirmed cases of COVID-19 across the world, with over 3.7 million deaths ^9^. Difficult to control viral transmission allied with the slow progress in the rollout of COVID-19 vaccines in most countries have contributed to the emergence of new variants of concern of SARS-CoV-2, which are more transmissible and can escape from natural and vaccine-acquired immunity ^10-13^.

SARS-CoV-2 is spread person to person mainly through exposure to respiratory fluids containing infectious virus. Virus exposure can occur in three main ways, which are not mutually exclusive: (i) inhalation of infectious virus present in very small fine droplets and aerosol particles; (ii) deposition of virus on exposed mucous membranes in the mouth, nose, or eye by direct splashes and sprays, and (iii) touching mucous membranes with hands contaminated by exhaled respiratory fluids containing virus or from touching fomites containing the virus^14^. Notably, SARS-CoV-2 has been found to have high person-to-person transmission through direct contact with infected individuals ^15^, especially by coughing, sneezing and even breathing/ talking by an infected person ^16-19^. SARS-CoV-2 enters the body through the mucous membranes of the eyes, mouth or nose and spreads to the nose line, sinus cavity, and throat until deposition into the human respiratory tract ^20^. Although transmission through direct contact, or airborne (respiratory droplets and/or aerosols) are considered to be the dominant routes for the spread of COVID-19^21, 22^, the transmission dynamics of SARS-CoV-2 by environmental surfaces and their role in the transmission chain remain unclear, and probably multifactorial. The risk of infection is influenced by the distance from the source, the amount of virus to which a person is exposed and the length of time since the virus has been deposited on the surface, since SARS-CoV-2 viability over time is influenced by environmental factors such as type of surfaces, temperature, humidity, and ultraviolet radiation (e.g., sunlight) ^21, 23, 24^.

Thus, understanding of distribution and patterns of environmental contamination by SARS-CoV-2 are relevant information for public health authorities. This knowledge allows the identification of critical points to establish effective control measures aimed at breaking transmission.

Several recent studies have investigated the presence of SARS-CoV-2 RNA in air and environmental surfaces, especially in health care settings ^25-33^. Previous studies under controlled laboratory conditions have demonstrated the ability of SARS-CoV-2 to remain infectious on different types of common surfaces, such as stainless steel, glass and paper, for up to 28 days at 20 °C ^34^, and it can also remain infectious in aerosols for up to 3 h ^35^. However, little is known about SARS-CoV-2 contamination of environmental surfaces in tropical public areas with a large flow and concentration of people. Therefore, studies investigating the presence of SARS-CoV-2 RNA on surfaces, and the infectious potential of these particles are of paramount importance.

To address this gap of knowledge, we investigated the presence of SARS-CoV-2 RNA on highly touched surfaces in Recife, a large city in Pernambuco state with a tropical monsoon climate. Samples were collected during the second wave of the COVID-19 in Brazil, one of the most severely affected countries by the pandemic. Our findings showed widespread viral contamination across many urban public settings and poor adherence to COVID-19 mitigation measures. Our data provide a real-world picture of SARS-CoV-2 dispersion in highly populated tropical areas and identify critical control points that need to be targeted to halt SARS-CoV-2 transmission.

## MATERIAL AND METHODS

### Study design and setting

This study was conducted in Recife, the capital of Pernambuco state, which is one of the most densely populated metropolitan regions in Brazil with 1,537,704 million people (https://cidades.ibge.gov.br/brasil/pe/recife). The city is located on the coast of Northeast coast of Brazil and has a tropical monsoon climate under the Köppen climate classification, with warm to hot temperatures and high relative humidity throughout the year.

This prospective cross-sectional study was designed busy areas and with a large flow and concentration of people. Initially, we subdivided Recife’s highly frequented places into 6 categories, including: a) transport terminals; b) health care units; c) public parks; d) public markets; e) beach areas; f) other public places (food supply center). A total of 400 environmental surface specimens were collected between Feb 2 and Feb 25, 2021 (Figure 1). Samples were collected between 9:00 a.m. and 1:00 p.m. During sample collection, the temperature was between 26°C to 32°C (average temperature 29°C) and the average humidity was 72%. Environment data was obtained from Time and Date AS website (http://www.timeanddate.com/weather/brazil/recife/climate). This coincided with a period of progressive increase in the number of COVID-19 cases in Pernambuco state and Brazil, representing the second wave of the COVID-19 pandemic in this part of the world (Figure 2) and also the beginning of COVID-19 vaccination efforts in this state. The ongoing pandemic of COVID-19 in the Pernambuco state has resulted in 499,572 laboratory-confirmed cases and 16,292 deaths as of 6 June 2021^36^. It is important to highlight that Recife has a high concentration of specialized hospitals and is considered a reference health center for the Northeast region of Brazil.

**Figure 1.**
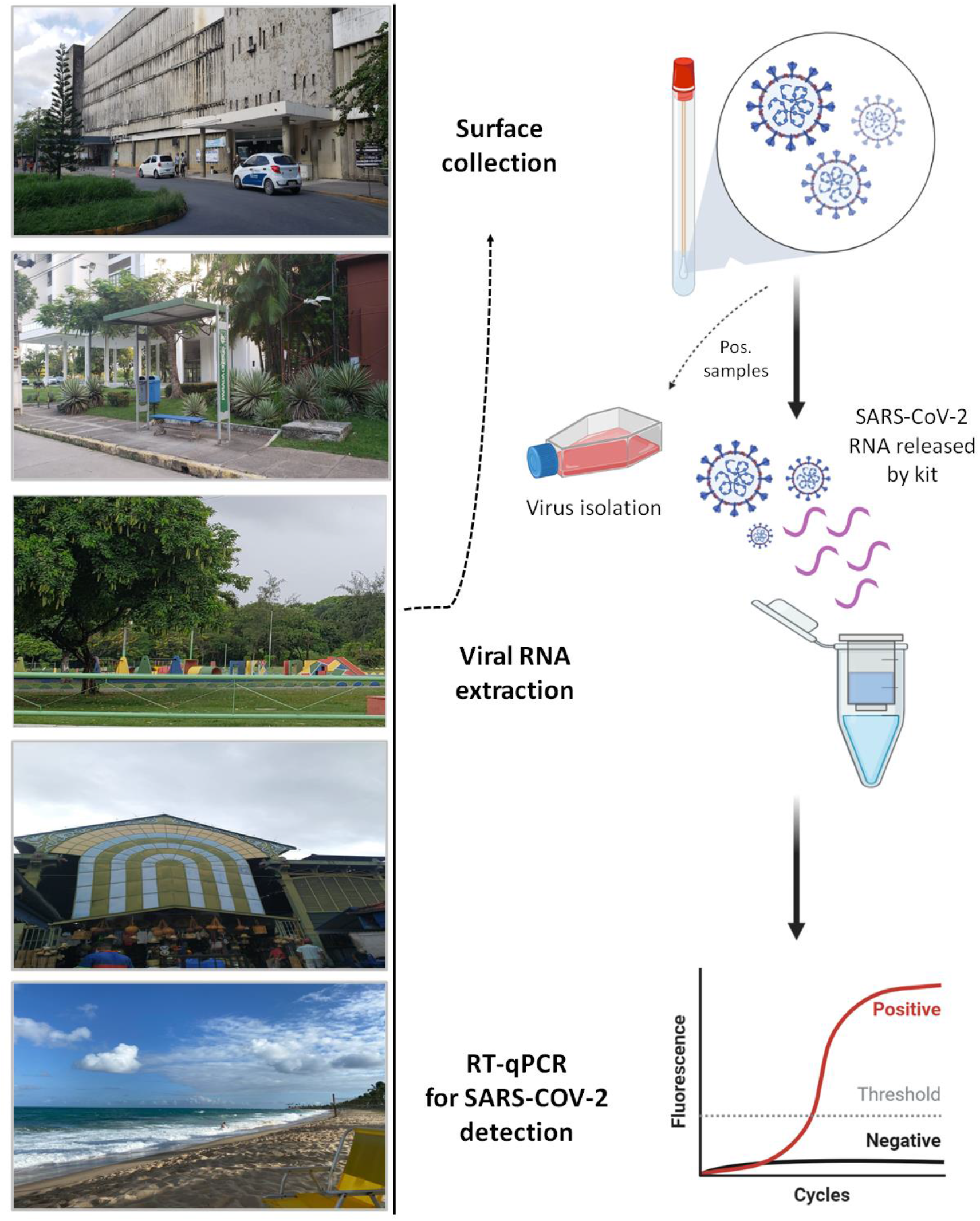
Study design showing the collection points of surface samples and the graphical workflow used to test the swabs. Created with Biorender.com

**Figure 2.**
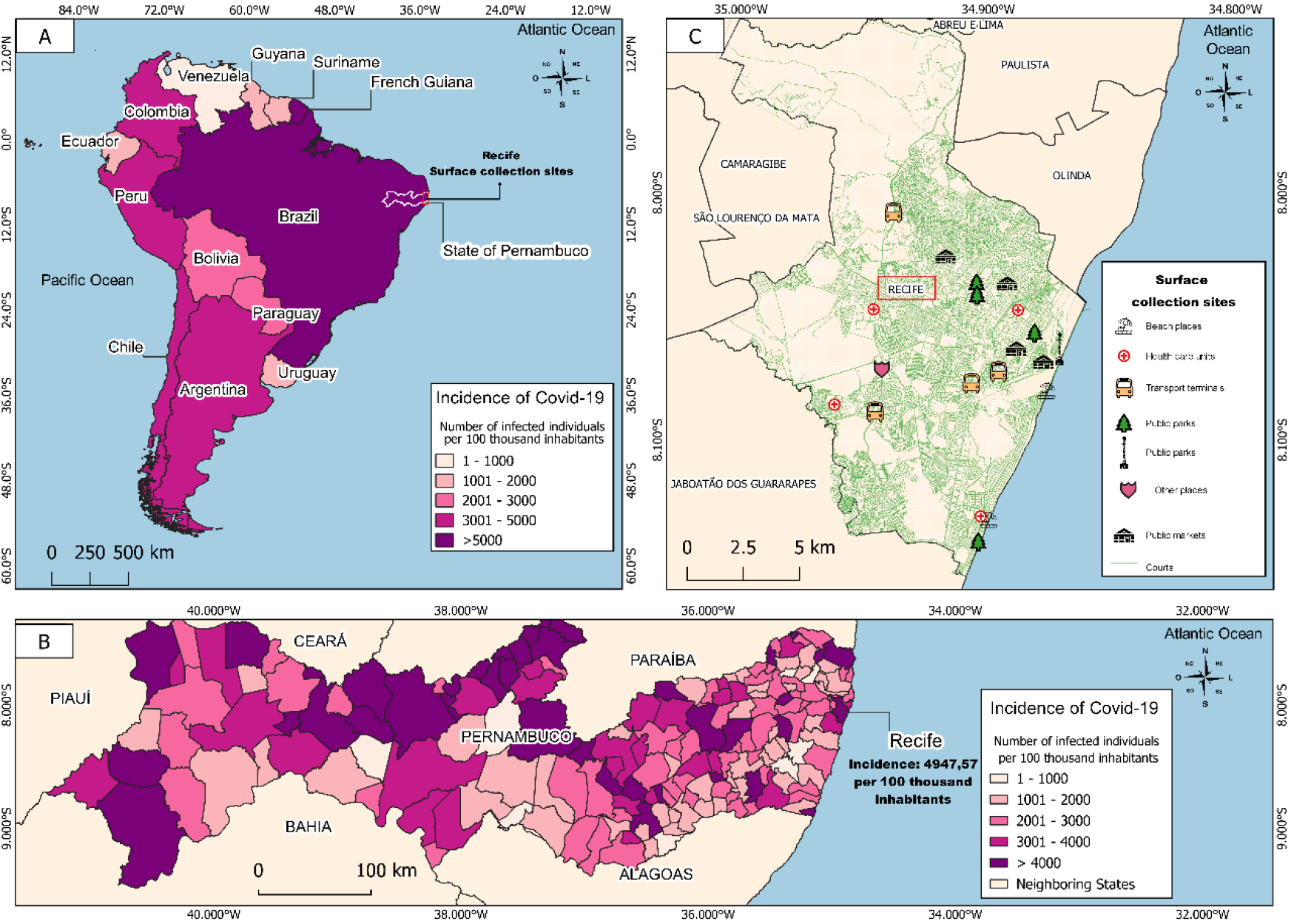
Spatial distribution of surface collection points and incidence of COVID-19 in Latin America and Pernambuco state, Brazil. Fig. 2A shows the incidence of COVID-19 per 100,000 inhabitants in Latin America. Fig. 2B shows the incidence of COVID-19 per 100,000 inhabitants in all cities in the state of Pernambuco, Northeast Brazil. Fig. 1C shows the spatial distribution of surface collection points (transport terminals, health care units, public parks, public markets, beach areas and other places) across Recife, Pernambuco state, Brazil.

### Sampling areas

#### Transport terminals

A total of 84 surface samples were collected from four public transport terminals with a large daily passenger flow and concentration. We strategically selected transport terminals that connect several cities in the metropolitan region of Recife. Twenty-one swabs were collected for each transport terminal. The collection points included the external area of the transport terminal and neighboring areas: (1) bus terminal entrance; (2) bus terminal exit; (3) bus terminal access; (4) subway station access; (5) ATM; (6) toilet; (7) handrail; (8) bench; (9) bus stop; (10) counter; (11) faucet; (12) ticket machine.

#### Health care units

A total of 84 surface samples were collected from four reference hospitals for treatment of COVID-19 patients in Recife, Brazil. Twenty-one swabs were collected for each hospital. The collection points included the external area of the hospital and neighboring areas: (1) principal entrance; (2) hospital access; (3) ambulatory entrance; (4) patient sample collection area; (5) toilet; (6) traffic light button; (7) coffee shop; (8) public phone; (9) bus stop; (10) resting area.

#### Public parks

A total of 105 surface samples were collected from five public parks. We strategically selected parks with high visitor flow, including children who access the playground. Twenty-one swabs were collected for each public park. The collection points included: (1) playground; (2) recreation area; (3) outdoor gym; (4) toilet; (5) handrail; (6) bus stop; (7) public bike station; (8) traffic light button; (9) coffee shop; (10) faucet.

#### Public markets

A total of 85 surface samples were collected from four public markets. Twenty-one swabs were collected for each public market with exception of one, where we collected twenty-two swabs. The collection points included: (1) principal entrance; (2) side entrance; (3) public market access; (4) toilet; (5) kiosk; (6) store; (7) food hall; (8) traffic light button; (9) faucet; (10) resting area; (11) outside area.

#### Beach areas

A total of 21 surface samples were collected from two beaches located in the coastal area of Recife, Brazil. Interestingly, the visited beaches had a high concentration of people during the time of surface collection and during all times of restrictive relaxation measures established by the state government during the COVID-19 pandemic. The collection points included: (1) toilets; (2) benches; (3) public bike station; (4) outdoor gym; (5) fresh green coconut; (6) handrails; (7) faucet; (8) traffic light button; (9) bus stop; (10) resting area.

#### Other areas

A total of 21 surface samples were collected from one food distribution center located in Recife, Brazil. We selected this place as it is a place which serves as a gateway for people from all over the Brazilian territory, and acts as a source of food supply for the Northeast of Brazil. The collection points included: (1) toilet; (2) restaurant; (3) handrail; (4) resting area.

### Surface sampling

Environmental samples were collected by qualified technicians who had received biosafety training and were equipped with personal protective equipment. For sample collection, sterile swabs (bioBoa Vista, Brazil) were used, that were put into a conical tube (15 mL) containing 2 mL of virus preservation solution (sterile phosphate-buffered saline, pH 7.2). Each swab was vigorously rubbed on the surface with a collection area of 25 cm^2^. Samples were collected from distinct types of materials, including metal, plastic, wood, rock, concrete, and glass. The time of collection and climate conditions of the day were recorded during sampling. In addition, an environmental site assessment questionnaire was applied to identify whether the collection environment and the population were following public health measures for preventing the rapid spread of SARS-CoV-2 and, subsequently, the COVID-19 transmission.

### Sample transfer and processing

Surface samples were collected and immediately stored at 4 °C prior to transfer to the biosafety level 3 laboratory (BSL-3) of Fiocruz Pernambuco, Brazil, where all samples were processed until 72 h after collection. After processing, each sample was taken directly tested according to the instructions described below.

### Viral RNA extraction and RT-qPCR for SARS-CoV-2 detection

Viral RNA was extracted from surface samples (140 μL of transport solution) using the QIAamp Viral RNA Mini Kit (QIAGEN, Germany) following the manufacturer’s protocol. RT-qPCR assay targeting the N protein according to protocols recommended by the Centers for Disease Control and Prevention - CDC USA, was used to detect SARS-CoV-2 (Supplementary Table 1) ^37^. Samples were considered positive when they presented amplification for N1 target, considering the threshold for cycle quantification (Cq) value of 40 ^37^. Samples with Cq ≥ 40 were considered as negative. Briefly, each reaction was prepared using the QuantiNova Probe RT-PCR Kit (QIAGEN, Valencia, CA, USA) following the manufacturer’s protocol and the CDC-USA recommendations in a total volume of 10 μL. Negative (extraction control and non-template control [NTC]) and positive controls (RNA extracted from SARS-CoV-2 cell supernatants) were included during all experiments. Primer and probe sequences were synthetized by IDT (Integrated DNA Technologies, Skokie, Illinois, USA). Thermal cycling was performed at 45 °C for 15 min for reverse transcription, followed by 95 °C for 5 min and then 45 cycles of 95 °C for 03 s and 55 °C for 30 s. All experiments were conducted using the Applied Biosystems QuantStudio 5 Real-Time PCR Systems (Applied Biosystems, USA). For data analysis, the QuantStudio software v1.5 was used with baseline and threshold automatic.

### Cells

African monkey green kidney-derived cell line Vero CCL-81 was used for virus isolation from positive environmental samples. Cells were cultured in Dulbecco’s modified Eagle’s medium (DMEM), high glucose (Gibco, USA) supplemented with 10% heat-inactivated fetal bovine serum, 100 U/ml penicillin and 100 μg/ml streptomycin (Gibco, USA); and maintained maintained in a humidified atmosphere, at 37 °C and 5% CO_2_.

### SARS-CoV-2 isolation

Vero CCL-81 cells were cultured in 12-well plates at a density of 2 × 10^5^ cells/well. After 24h, the culture media was removed and cells were incubated with 300 µL of undiluted and filtered surface samples at 37°C, 5% CO_2_, for 1h. Fresh media supplemented with 2% FBS (700 µL) was added to the cells and they were maintained at 37°C, 5% CO^2^. Cells were monitored daily for the visualization of virus-induced cytopathic effect (CPE). CPE images were acquired in Carl Zeiss Axio Observer 5 microscope coupled to a photographic camera. After 3 days post infection (d.p.i.) supernatants were collected and 300 µL were transferred to a new 12-well plate. This procedure was repeated until completing three passages (P1, P2 and P3). Following this, cell culture supernatants were collected on t=0h and t=72h in each passage for viral RNA extraction and possible SARS-CoV-2 detection by RT-qPCR. All experiments were performed in a BSL-3 facility.

### Environmental site assessment questionnaire

Data regarding the social distancing, mask wearing, availability of hand sanitizers and COVID-19 control measures during sample collection in all locations was obtained using a structured questionnaire following the recommendations and guidelines established by WHO and CDC ^38^. The questions aimed to identify the implementation and compliance with COVID-19 prevention measures, including social distancing, mask wearing, the availability of hand sanitizers, body temperature measurements for screening and the presence of informative charts for people education. The questionnaires were made with qualitative, with “yes” or “no” input, or quantitative inquiries.

### Spatial location of collection surfaces

To georeference the locations where surface samples were obtained, we used the QGIS software (https://qgis.org/en/site/) to generate a map using the geographic coordinates of each publicly available location at https://www.google.com.br/maps. First, we created a graduate map with information about the incidence of COVID-19 in the countries of Latin America (Figure 2A) and all cities located in the State of Pernambuco, Brazil (Figure 2B). The incidence per 100 thousand inhabitants was calculated using the database of the last Brazilian census available at http://censo2010.ibge.gov.br and epidemiological reports of COVID-19 cases from the Pernambuco State Health Department ^36^ and the World Organization Health (WHO)^39^. Furthermore, we showed the spatial distribution of urban public places where the samples were collected including transport terminals, health care units, public parks, public markets, beach areas, and other areas. We acquired the cartographic base in shapefile format through the Brazilian Institute of Geography and Statistics (IBGE) in the Geocentric Reference System for the Americas (SIRGAS) 2000 (Figure 2C).

### Data analysis

GraphPad Prism software version 5.01 for Windows (GraphPad Software, La Jolla, California, USA) was used to plot most graphics. The association analysis between collection locations and type of materials was demonstrated based on the results from 97 positive surfaces collected in this study using the web-based Circos table viewer, version 0.63-9 (https://www.mkweb.bcgsc.ca/tableviewer/visualize/) ^40^.

### Ethics approval

This study was reviewed and approved under protocol number 03/2021 by the Fiocruz Pernambuco Internal Biosafety Commission, as part of quality assurance for working with highly pathogenic virus.

## RESULTS

### Distribution of surface samples according to collection area and type of material

A total of 400 surface samples were collected in Recife, Pernambuco state in 19 sites divided into 6 subgroups (health care units, transport terminals, public parks, public markets, beach areas, and a food distribution center). A total of 97 surface samples (24.2%) tested positive for SARS-CoV-2 RNA using the CDC-USA protocol by RT-qPCR (Figure 3a, Supplementary Table 1) in 18 out of 19 sites sampled (Supplementary Table 2). The only site that tested negative was a public market. SARS-CoV-2 RNA was detected in 47 (48.4%) surface samples collected around transport terminals, followed by health care units (26/97, 26.8%), public parks (14/97, 14.4%), public markets (4/97, 4.1%), beach areas (4/97, 4.1%), and other places (2/2.0%) (Figure 3b, Supplementary Table 3). Regarding the type of material where environmental samples were collected, SARS-CoV-2 RNA was found most frequently on metal (45/97, 46.3%), followed by plastic (18/97, 18.5%), wood (12/97, 12.3%), rock (10/97, 10.3%), concrete (8/97, 8.2%), glass (2/97, 2.0%), and other (ceramic and rubber) (2/97, 2.0%) (Figure 3c). Positive samples were predominantly found in toilets, ATMs, handrails, playground, and outdoor gym; highlighting the importance of these fomites in SARS-CoV-2 surface contamination.

**Figure 3.**
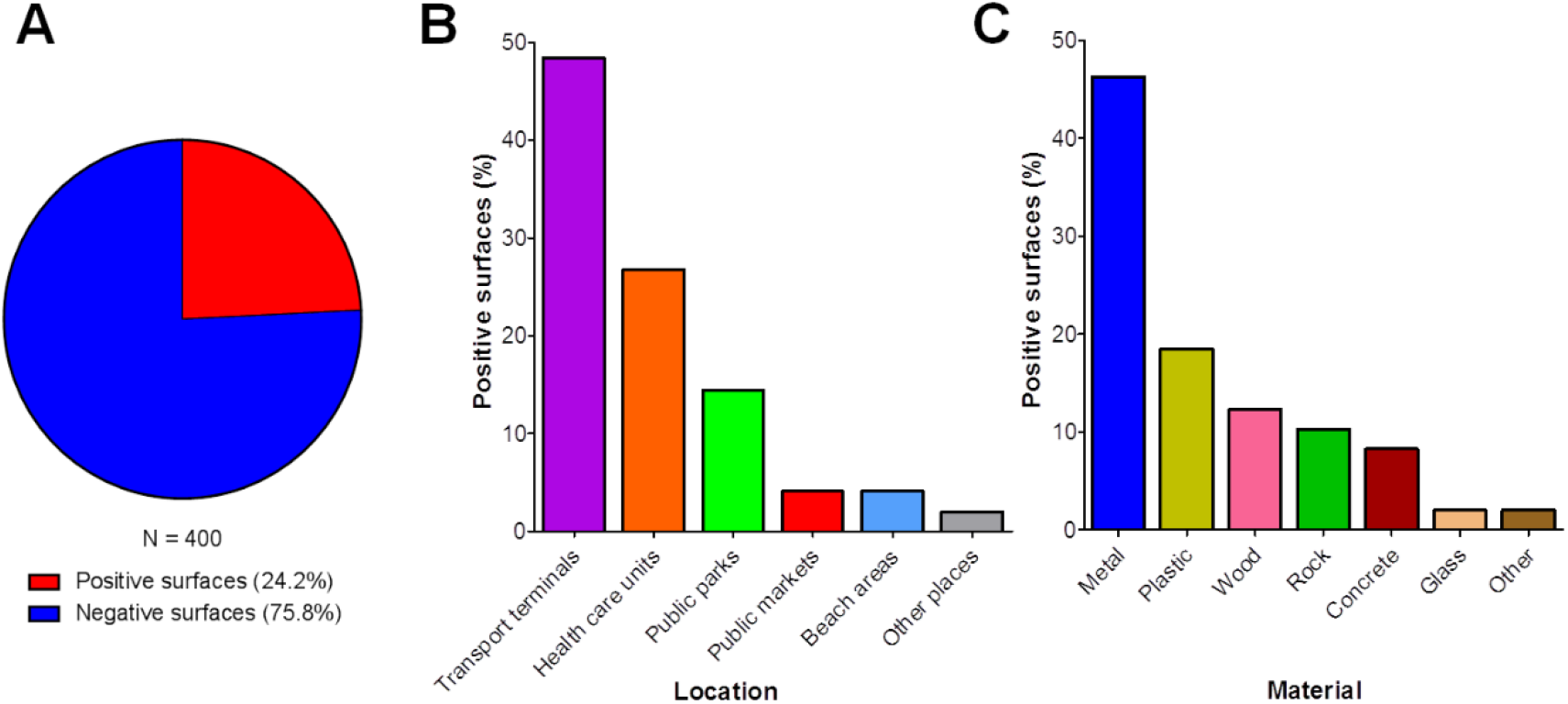
Overall results for SARS-CoV-2 detection in surface samples. Fig. 3A shows the distribution of positive and negative samples using a total of 400 environmental samples. Fig. 3B shows the distribution of positive samples according to the collection areas; including transport terminals, health care units, public parks, public markets, beach areas, and other places Fig. 3C shows the distribution of positive samples according to the type of material including metal, plastic, wood, rock, concrete, glass and other.

### Distribution of positive surface samples according to point of collection

#### Transport terminals

Forty-seven (48.4%) surface samples were positive for SARS-CoV-2 RNA around public transport terminals with Cq values ranging from 31.1 to 38.7 by RT-qPCR (Supplementary Table 3). Positive samples were distributed particularly in eleven different locations, including ATM (9/47, 19.1%), handrails (9/47, 19.1%), bus terminal access (7/47, 14.8%), bench (6/47, 12.7%), toilet (5/47, 10.6%), ticket machine (3/47, 6.3%), bus stop (2/47, 4.2%), subway station access (2/47, 4.2%), faucet (2/47, 4.2%), bus terminal exit (1/47, 2.1%), and ticket counter (1/47, 2.1%) (Figure 4a, Supplementary Table 3).

**Figure 4.**
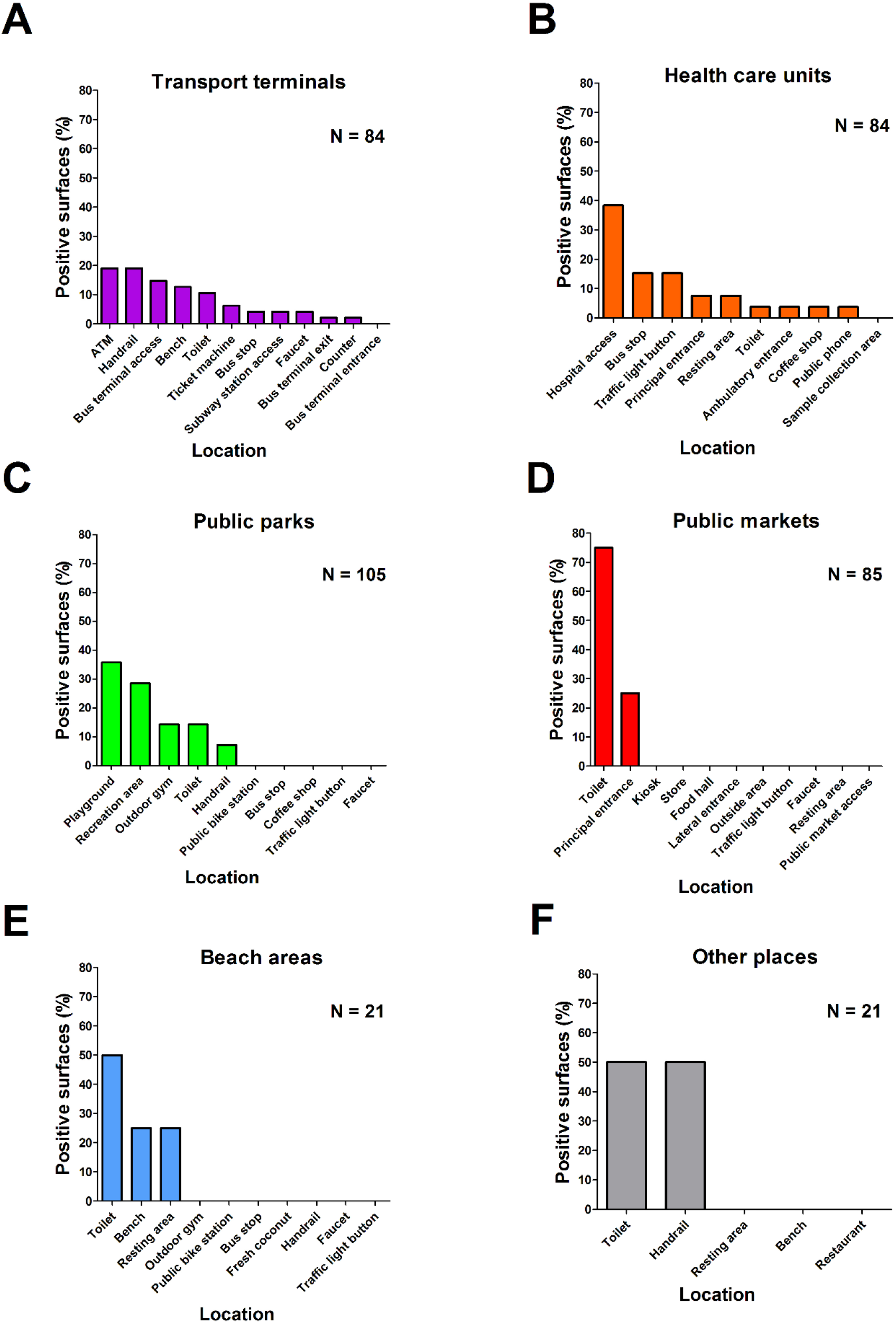
Distribution of positive surface samples according to collection areas. Fig. 4A shows the distribution of positive samples around transport terminals. Fig. 4B shows the distribution of positive samples around health care units. Fig. 4C shows the distribution of positive samples around public parks. Fig. 4D shows the distribution of positive samples around public markets. Fig. 4E shows the distribution of positive samples around beach areas. Fig. 4F shows the distribution of positive samples around the other areas (including one food distribution center).

#### Health care units

Twenty-six (26.8%) surface samples were positive for SARS-CoV-2 RNA in the surroundings of health care units with Cq values ranging from 31.1 to 38.7 by RT-qPCR (Supplementary Table 3). Positive samples were found in nine different locations from four reference hospitals for COVID-19 treatment. The areas with highest number of positive samples were hospital access (10/26, 38.4%), bus stop (4/26, 15.3%), traffic light button (4/26, 15.3%), principal entrance (2/26, 7.6%), resting area (2/26, 7.6%), toilet (1/26, 3.8%), ambulatory entrance (1/26, 3.8%), coffee shop (1/26, 3.8%), and public phone (1/26, 3.8%) (Figure 4b, Supplementary Table 3).

#### Public parks

Fourteen (14.4%) surface samples were positive for SARS-CoV-2 RNA around public parks, with Cq values ranging from 36.2 to 39.7 by RT-qPCR (Supplementary Table 3). Positive samples were collected from five different locations, including playground (5/14, 35.7%), recreation area (4/14, 28.5%), outdoor gym (2/14, 14.2%), toilet (2/14, 14.2%), and handrails (1/14, 7.1%) (Figure 4c, Supplementary Table 3). There were no positive samples from the public bike station, bus stop, coffee shop, traffic light button, or faucet.

#### Public markets

Three out of four public markets sampled returned at least one positive sample. Four (4.1%) surface samples were positive for SARS-CoV-2 RNA in public markets with Cq values ranging from 36.9 to 38.1 by RT-qPCR (Supplementary Table 3). Positive samples were collected from two different locations, including toilets (3/4, 75.0%) and principal entrance (1/4, 25.0%) (Figure 4d, Supplementary Table 3). No positive samples were found at the kiosk, store, lateral entrance, outside area, food hall, public market access, traffic light button, faucet, or resting area.

#### Beach areas

Four (4.1%) surface samples were positive for SARS-CoV-2 RNA in beach areas with Cq values ranging from 36.1 to 37.9 by RT-qPCR (Supplementary Table 3). Positive samples were collected from three different locations, including toilets (2/4, 50.0%), bench (1/4, 25.0%), and resting area (1/4, 25.0%) (Figure 4e, Supplementary Table 3). No positive samples were detected from the outdoor gym, public bike station, bus stop, fresh coconut, handrail, faucet, or traffic light button.

#### Other places

Two (2.0%) surface samples were positive for SARS-CoV-2 RNA around one food distribution center with Cq values ranging from 38.0 to 38.7 by RT-qPCR (Supplementary Table 3). Positive samples were collected from two different locations, including toilet (1/2, 50.0%) and handrails (1/2, 50.0%) (Figure 4f, Supplementary Table 3). No positive samples were found in restaurants or resting benches.

### Types of surface materials positive for SARS-CoV-2 RNA

From the 47 positive samples in transport terminals, 21 (44.6%) samples were identified mainly on metal surfaces, especially from handrails at bus terminals, ATM button, protection grid, and faucet. 19 (19.1%) samples were recovered from plastic surfaces, especially around biometrics sensors in ATMs and faucets in the toilet. 5 (10.6%) samples were found in concrete surfaces, most being found in pillars near the bus stop and one sampled from a bench. Four (8.5%) samples were collected on rock surfaces, with virus being detected on walls in the toilet and bus terminal, and one sample was collected at the terminal service desk. Four (8.1%) samples were identified on wood surfaces, all being from benches near the bus stop of transport terminals. Two (4.2%) samples were detected on glass surfaces, mainly on the ticket machine screens. In addition, one (2.1%) sample was collected on a toilet seat (porcelain) and one (2.1%) was detected on the ticket machine (rubber) (Figure 5, Supplementary Table 3).

**Figure 5.**
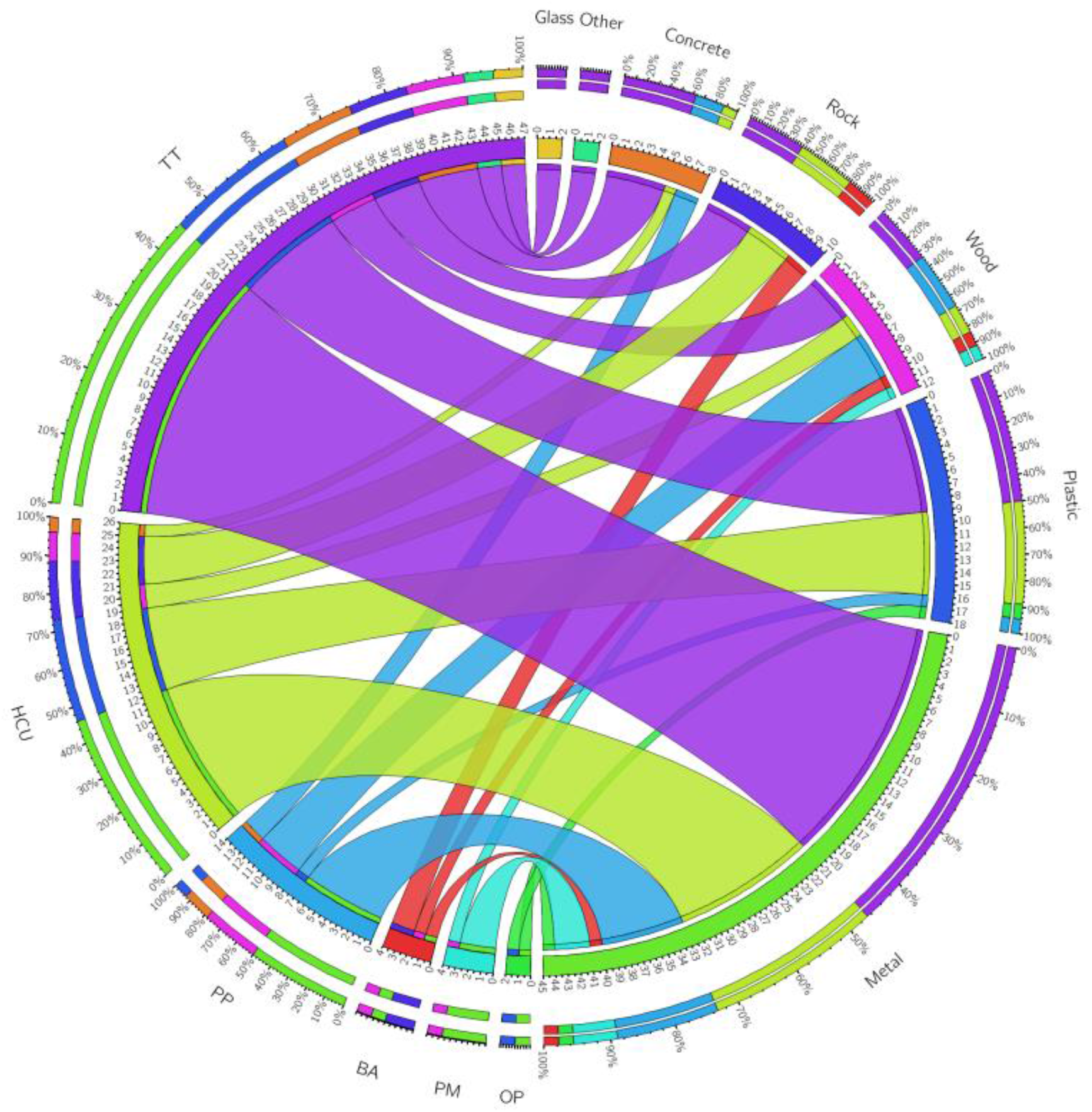
Association between the surface collection areas, and type of material where SARS-CoV-2 RNA was detected. TT: transport terminals; HCU: health care units; PP: public parks; PM: public markets; BA: beach areas; OP: other places.

From the 26 positive samples found in health care units and neighboring areas, 12 (46.1%) samples were recovered from metal surfaces mostly, located at the entrance to hospitals and near bus stops. 7 (26.9%) samples were identified in plastic surfaces, especially from traffic light buttons, near bus stops, and in the toilets. Four (15.3%) samples were detected in rock surfaces found at the entrance to hospitals. Two (7.6%) samples were identified in wood surfaces at the entrance to hospitals. One (3.8%) sample was detected on the concrete surface from a nearby bus stop (Figure 5, Supplementary Table 3).

From the 14 positive samples found in public parks, seven (50.0%) samples were identified on the metal surfaces of handrails in the playground and outdoor gym. Four (28.5%) samples were recovered from wood surfaces in the playground, and one tourist attraction point. Two (14.2%) samples were detected in concrete surfaces of the playground. One (7.1%) sample was identified in plastic surface from a faucet in the toilet (Figure 5, Supplementary Table 3).

From the four positive samples in public markets, three (75.0%) samples were detected on metal surfaces at the entrance to public markets, and from a toilet faucet. One (25.0%) sample was detected identified in wood surfaces from a door in the toilet.

From the four positive samples in beach areas, two (50.0%) were detected in rock surfaces, one from toilet wall and one from a bench. One (25.0%) sample was identified in a metal surface from a faucet in the toilet, and a further one (25.0%) was detected in a wood surface on a handrail that gives access to the beach.

Lastly, of the two positive samples from two food distribution center, one (50.0%) sample was detected on a plastic surface from a faucet in the toilet and one (50.0%) was identified on a metal handrail surface at the entrance of a bank (Figure 5, Supplementary Table 3).

### Viability of SARS-CoV-2 from positive surfaces samples

To assess infectivity of samples that tested positive by RT-qPCR, nine samples with Cq value <34 (Cq ranging from 31.0 to 33.7) were inoculated into 12-well plates seeded with Vero CCL-81 cells. Samples were considered negative after three blind passages of the supernatant. Under these conditions it was not possible to isolate the virus, as determined by the absence of CPE and negative RT-qPCR results from third passage supernatant (Supplementary Figure 2, Supplementary Table 4). The risk of infection from these contaminated surfaces is therefore not clear.

### Poor adherence of COVID-19 mitigation measures by society

Data regarding the adoption of public health measures, and community perception of COVID-19 disease was collected during surface collection in all locations by using a structured environmental site assessment questionnaire. In the 19 collection points, 70% alcohol-based hand sanitizer was available at the entrance in 26.3% (5/19) of the locations, whereas 42.1% (8/19) had a sink with soap and water for hand hygiene. Temperature measurements at the entrance was carried out in 15.8% (3/19) of the sites, and information material on preventive measures to prevent SARS-CoV-2 transmission was found in 42.1% (8/19) of the sites. High mask wear adherence was seen (94.7% [18/19]), although only 57.3% of people (average calculated for every 10 people per collection point) were wearing masks in a proper way. Regarding social distancing, only 26.3% (5/19) of the people present at collection points were maintaining the recommended social distance of 2 m. Furthermore, only 5.3% (1/19) of collection sites were limiting the number of people who accessed the location point (Table 1). We found no positive correlation between adherence of COVID-19 mitigation measures and SARS-CoV-2 positivity (data not shown). Overall, our findings indicated poor adherence of COVID-19 mitigation measures in our study areas.

**Table 1.**
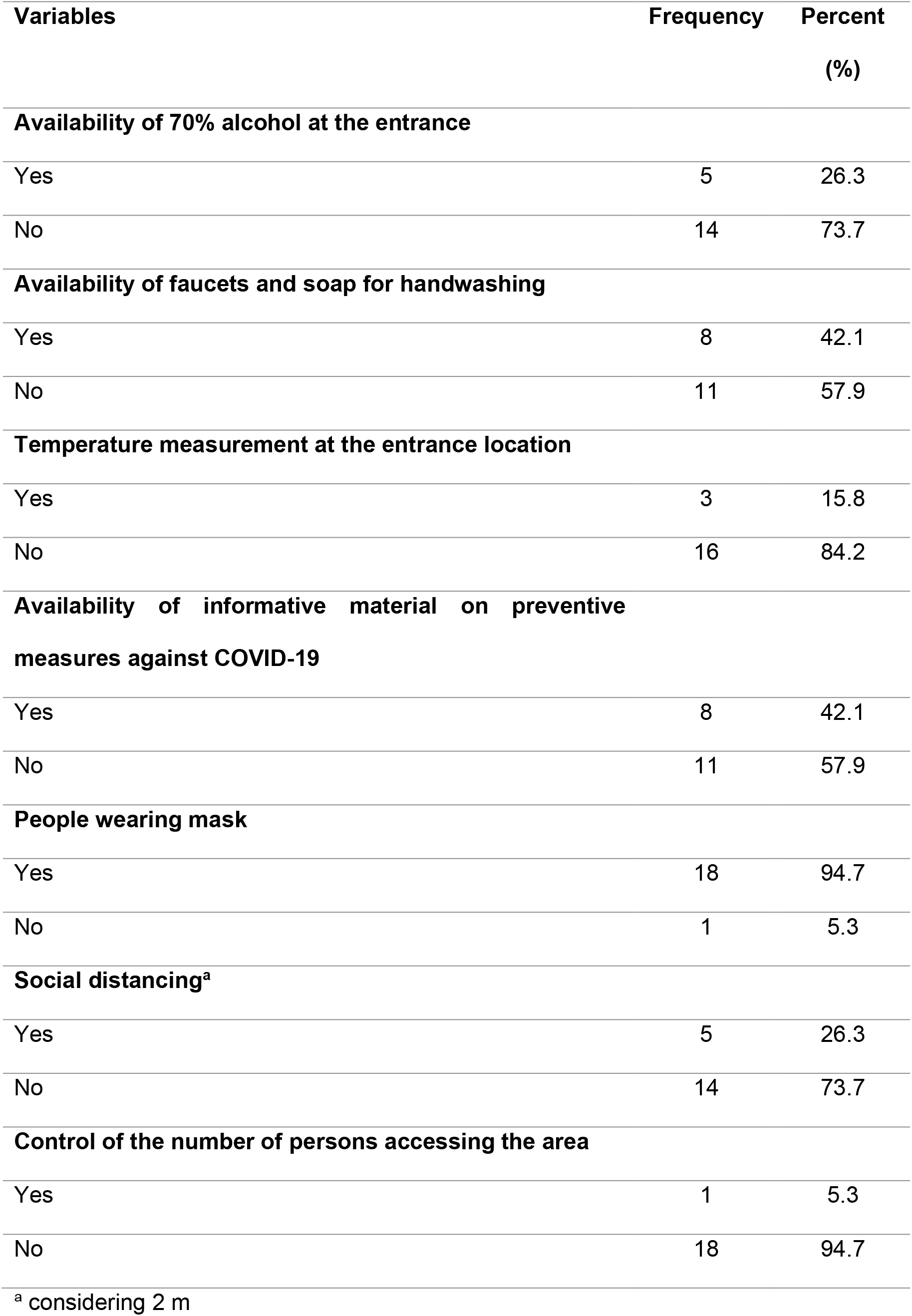
Evaluation of safety procedure protocol implementation against COVID-19 at collection areas (n=19).

| **Variables** | **Frequency** | **Percent (%)** |
| --- | --- | --- |
| **Availability of 70% alcohol at the entrance** |  |  |
| Yes | 5 | 26.3 |
| No | 14 | 73.7 |
| **Availability of faucets and soap for handwashing** |  |  |
| Yes | 8 | 42.1 |
| No | 11 | 57.9 |
| **Temperature measurement at the entrance location** |  |  |
| Yes | 3 | 15.8 |
| No | 16 | 84.2 |
| **Availability of informative material on preventive measures against COVID-19** |  |  |
| Yes | 8 | 42.1 |
| No | 11 | 57.9 |
| **People wearing mask** |  |  |
| Yes | 18 | 94.7 |
| No | 1 | 5.3 |
| **Social distancing<sup>a</sup>** |  |  |
| Yes | 5 | 26.3 |
| No | 14 | 73.7 |
| **Control of the number of persons accessing the area** |  |  |
| Yes | 1 | 5.3 |
| No | 18 | 94.7 |

<sup>a</sup> considering 2 m

## DISCUSSION

Since the emergence of SARS-CoV-2, first identified in China, the highly pathogenic coronavirus has spread rapidly around the world causing an unprecedented health security crisis and drastically affecting the global economic stability. Thus, understanding the modes of transmission of SARS-CoV-2 among humans is a critical step to establish effective prevention policies and prioritize resources to break the chain of SAR-CoV-2 transmission. The transmission through direct contact and via airborne (respiratory droplets and/or aerosols) are pointed as the dominant routes for the transmission of SARS-CoV-2 in humans ^21, 22, 41^ and animal models, like ferrets ^42^, golden hamsters ^43^, and mices^44^. Similarly, many studies conducted on the spread of other respiratory viruses, including influenza virus ^45, 46^, respiratory syncytial virus (RSV) ^47^, and severe acute respiratory syndrome coronavirus (SARS-CoV-1) ^48^, evidenced that these respiratory viruses can be exhaled and transmitted via airborne. However, the transmission dynamics of SARS-CoV-2 by environmental surfaces and their role in the transmission chain remains unclear and may be multifactorial, especially in urban areas with a large flow and concentrations of people with real-life challenges. Here, we investigated the presence of SARS-CoV-2 RNA on public high-touch surfaces in a large metropolitan city during the second wave of the COVID-19 pandemic in Brazil.

A recent study investigated the presence of SARS-CoV-2 RNA on public surfaces in Belo Horizonte, a large city with a tropical savanna climate in Southeast Brazil. A total of 933 swabs collected from different locations including health care units, public squares, bus terminals, public markets, and other public places between April and June 2020^49^. The results showed that 49 (5.25%) of surface samples were tested positive for SARS-CoV-2 RNA, although the infectious potential of positive samples was not investigated. Considering the proportion of positivity in the different places, the authors pointed out that bus terminals exhibited the highest positivity rate, followed by public markets, public squares, and health care units ^49^. In our study, we found higher positivity of SARS-CoV-2 RNA (97/400, 24.2%) detection of surfaces compared to the Belo Horizonte survey. In our study, most of the positive samples in our study were detected in the surroundings of transport terminals areas (48.4%), followed by health care units (26.8%), public parks (14.4%), public markets (4.1%), and beach areas (4.1%). The difference in the positivity rate of both cities cannot be explained by climate differences as Recife is hotter and more humid than Belo Horizonte, conditions that decreases the stability of SARS-CoV-2 in the environment ^50^ and its transmissibility^51^. A more plausible explanation for this disparity is the number of confirmed COVID-19 cases in these cities by the time of sample collection. Whereas Belo Horizonte registered 400 to 5,000 (https://ciis.fmrp.usp.br/covid19/bh-mg/) daily cases between April and June 2020, Recife had 60,000 to 70,000 (https://ciis.fmrp.usp.br/covid19/recife-pe/) in February 2021. Taken together, our findings are in agreement with others and indicates widespread SARS-CoV-2 surface contamination in public urban places with a large flow of people ^49, 52^.

Regarding the distribution of positive samples according to the type of material, we found the SARS-CoV-2 RNA mainly on metal, followed by plastic, wood, rock, concrete, and glass. Similarly, a recent urban study found the SARS-CoV-2 RNA on different types of materials, the majority on metal, concrete, rock, brickwork, wood, and glass^49^. Interestingly, our data demonstrated that the positive samples for SARS-CoV-2 RNA were mainly collected in toilets. These findings also corroborate data obtained by other groups^26, 30, 53^, which toilets as an area of high positivity rate for SARS-CoV-2 RNA. Additionally, our findings revealed other specific locations with high rates of positivity: ATMs, handrails, playgrounds, and outdoor gyms.

Previous studies performed under controlled laboratory conditions have shown that SARS-CoV-2 remains infectious on different types of surfaces, such as stainless steel, glass and paper, for up to 28 days at 20 °C^34^, depending on type of environmental surface; and can remain viable in aerosols for up to 3 h ^35^. Notably, the viral load decreases over time and depends on the length of time since the virus has been deposited on the surface, which may be reflected in the presence of infectious or non-infectious viral particles and, consequently, infection risk in humans ^21, 23, 24^. Another important factor that must be considered is the minimal infectious dose of SARS-CoV-2 to start an effective infection in humans, which has not yet been clarified. In order to elucidate the transmission dynamics of SARS-CoV-2 by environmental surfaces in real-life conditions, several studies have investigated the presence of SARS-CoV-2 in air and environmental surfaces/areas, including health care settings ^25-31, 33^ and urban settings^49, 52-55^. In general, these studies have found varying levels of environmental contamination, ranging from extensive ^25, 26^ to low contamination ^31, 49^, or even no contamination of SARS-CoV-2 RNA. However, many of these studies did not determine the ability of SARS-CoV-2 to be cultured from such environmental swabs, which would help to understand the implications of SARS-CoV-2 RNA positive environmental samples in terms of infectious potential for the human population ^25, 27^. In this study, we evaluated the infectious potential of positive surface samples (Cq value <34) in Vero CCL-81 cells, but SARS-CoV-2 could not be cultured. This finding is supported by recent studies, which have demonstrated the low potential infectious from the environmental swabs using cell culture ^25, 31, 56^. This may explain the lack of success in virus isolation given the short half-life of SARS-CoV-2 in the environment. Serial sampling of highly touched surfaces in places with large people flow might produce culturable SARS-CoV-2. Nevertheless, our findings identify the locations and objects that pose the highest risk of contamination through fomites and should be considered as COVID-19 critical control points. The difficulty in culturing viruses from environmental samples arises from low viral load concentrations and instability of SARS-CoV-2 outside the human host. Recent studies aggregated environmental sampling has shown high RT-qPCR Cq values (>30) for most of the positive samples, which may explain the difficulty of SARS-CoV-2 to be cultured from the environmental specimens ^25, 33,49^. Other studies have suggested that several environmental stressors can compromise and damage the integrity of SARS-CoV-2 viral particles, including temperature and relative humidity ^34, 50^.

SARS-CoV-2 contamination of public surfaces suggests the circulation of infected people and the risk of infection in these locations either by direct or indirect contact with infected patients. Direct contact with an infectious source is important for the establishment of COVID-19 clinical features and this has been established using animal models. Transmission studies in the ferret SARS-CoV-2 model have demonstrated that airborne transmission is likely but is considerably less efficient than direct contact transmission, whereby direct contacting animals are exposed to infected ferrets and share with them the same food, water, bedding, and breathe the same air^42, 57^.

Regarding the adherence of COVID-19 mitigation measures by society, a number of studies have been performed in order to evaluate the adoption of measures to prevent the SARS-CoV-2 transmission ^58-60^. To assess the community’s adherence to mitigation measures to combat the rapid spread of SARS-CoV-2, a recent cross-sectional study conducted in Malaysia employed 4,850 Malaysian residents, between 27th March and 3rd April 2020 ^59^. The findings revealed that most participants (83.1%) held positive attitudes toward the successful control of COVID-19, the capacity of Malaysia to counter rapid spread of the disease (95.9%) and the way the Malaysian government was facing the COVID-19 crisis (89.9%). Furthermore, most participants were also taking precautions such as practicing hand hygiene (87.8%) and avoiding large gatherings (83.4%) ^59^. Interestingly, the number of COVID-19 cases in Malaysia remained stable, with a progressive increase observed only between September and November 2020 (https://ourworldindata.org/covid-cases). In contrast, a community-based cross-sectional study done in Northeast Ethiopia, evaluated the adherence towards COVID-19 mitigation strategies by society among 635 individuals from April 20–27, 2020 ^58^. The results showed that approximately half of the study participants had poor adherence towards COVID-19 mitigation measures. In the current analysis, although the number of places evaluated was limited (19), it is important to highlight that these are places with a high flow and concentration of people. Our data demonstrated low adherence of COVID-19 mitigation measures by society regarding the social distancing, effective use of masks, precaution measures adoption and community’s perception about the COVID-19 disease. Taken together, these results highlight the importance of consistent messaging from government and health authorities to improve levels the adoption of measures to prevent and contain the spread of SARS-CoV-2.

In summary, our data demonstrated the extensive viral RNA contamination of surfaces in a range of public urban settings in the absence of viral isolation, which suggests low potential risk from environmental contamination for the human population. However, we identified poor adherence to COVID-19 mitigation policies by wider society regarding the adoption of control measures, and this may be reflected in the frequent detection of the viral RNA. Studies such as these can contribute to assess the prevalence of SARS-CoV-2 in specific settings. Finally, we suggest that further studies are urgently performed to elucidate the relative contribution of various modes of transmission for SARS-CoV-2 in both healthcare and urban-settings.

## Data Availability

All data is provided in the manuscript and in the supplementary files

## Acknowledgments

We are grateful to Msc. Larissa Krokovsky for her support and critical analysis for the design of the work. We thank Josenildo do Sobral for helping to collect surface samples. We would like to express our sincere gratitude and appreciation to the Fiocruz Pernambuco team for the support.

## Authorship contribution statement

L.P., A.K. and S.J.R.d.S. conceived the work. S.J.R.d.S., J.C.F.N., W.P.M.d.S.R., C.T.A.S., P.G.S., R.P.G.M., A.A.M., B.N.R.S. and J.J.F.M. performed the experiments. S.J.R.d.S., J.C.F.N., W.P.M.d.S.R., P.G.S. A.A.M., J.J.F.M., A.K. and L.P. performed data analysis and interpretation. S.J.R.d.S., J.C.F.N., W.P.M.d.S.R., A.A.M. and J.J.F.M. wrote the original draft. S.J.R.d.S., A.K. and L.P. wrote the final manuscript. LP supervised the work. All authors critically revised the manuscript and approved the final version of the submitted manuscript.

