## Supplementary figures for "Widespread Contamination of SARS-CoV-2 on Highly Touched Surfaces in Brazil During the Second Wave of the COVID-19 Pandemic"

**Figure S1.** Standard curve using N1 and N2 primers to detect SARS-CoV-2 RNA extracted from cell supernatants.

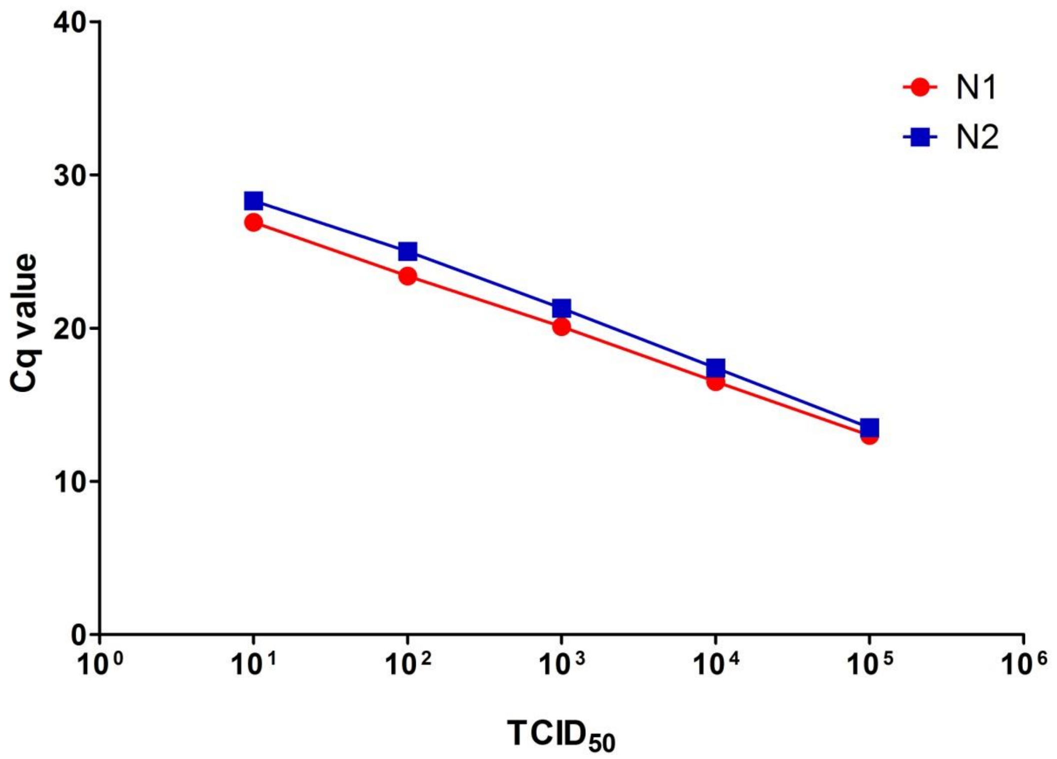


$TCID_{50}$: 50% Tissue Culture Infectious Dose

Cq: Cycle quantification

**Figure S2.** Attempted to SARS-CoV-2 isolation using the Vero CCL-81 cell lineage.

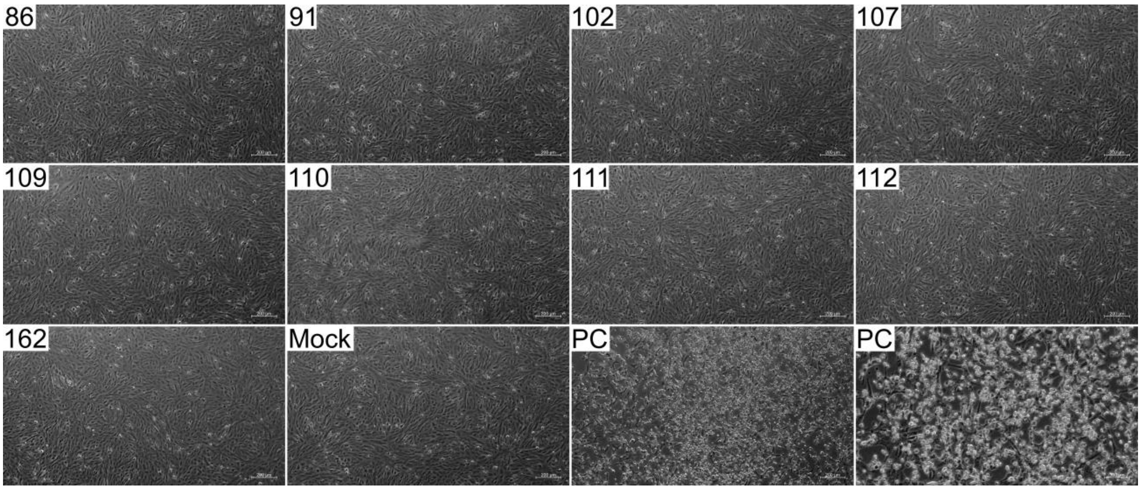


**Cytopathic effect observed in positive surface samples.** Specimens with a lower Cq value (86, 91, 102, 107, 109, 110, 111, 112 and 162) among samples tested positive for SARS-CoV-2 after three passages. Mock: uninfected cells; PC: positive control (SARS-CoV-2 isolated from patient sample).

**Table S1.** Primers used in this study for SARS-CoV-2 detection.

| Target | Primer name | Oligonucleotide Sequence (5'-3')[a] | Assay |
|---|---|---|---|
| N | 2019-nCoV_N1-F | GACCCCAAAATCAGCGAAAT | RT-qPCR |
| | 2019-nCoV_N1-R | TCTGGTTACTGCCAGTTGAATCTG | RT-qPCR |
| | 2019-nCoV_N1-P | FAM-ACCCCGCATTACGTTTGGTGGACC-BHQ1 | RT-qPCR |
| | 2019-nCoV_N2-F | TTACAAACATTGGCCGCAAA | RT-qPCR |
| | 2019-nCoV_N2-R | GCGCGACATTCCGAAGAA | RT-qPCR |
| | 2019-nCoV_N2-P | FAM-ACAATTTGCCCCCAGCGCTTCAG-BHQ1 | RT-qPCR |

[a] Primers were designed by CDC-USA.

**Table S2. Positivity of SARS-CoV-2 RNA on public touched surfaces at different locations in Recife, Pernambuco state, Brazil.**

| **Location** | **Site** | **Number of samples collected** | **Number os positive surfaces (%)** |
|---|---|---|---|
| Transport terminals | 01 | 21 | 20 (95.2%) |
| | 02 | 21 | 16 (76.1%) |
| | 03 | 21 | 8 (38.0%) |
| | 04 | 21 | 3 (14.2%) |
| Health care units | 05 | 21 | 4 (19.0%) |
| | 06 | 21 | 3 (14.2%) |
| | 07 | 21 | 18 (85.7%) |
| | 08 | 21 | 1 (4.76%) |
| Public parks | 09 | 21 | 3 (14.2%) |
| | 10 | 21 | 4 (19.0%) |
| | 11 | 21 | 3 (14.2%) |
| | 12 | 21 | 2 (9.52%) |
| | 13 | 21 | 2 (9.52%) |
| Public markets | 14 | 21 | 2 (9.52%) |
| | 15 | 21 | 0 (0%) |
| | 16 | 21 | 1 (4.76%) |
| | 17 | 22 | 1 (4.54%) |
| Beach places | 18 | 21 | 4 (19.0%) |
| Other places | 19 | 21 | 2 (9.52%) |

**Table S2.** Positive samples for SARS-CoV-2 RNA on public touched surfaces at different locations in Recife, Pernambuco state, Brazil.

| Location | Sample ID* | Surface | Specific location | Type of material | Cq value (N1) |
|---|---|---|---|---|---|
| **Health care units** | 01 | Standing rest bar | Ambulatory entrance | Metal | 37.8 |
| | 08 | Handle | Hospital access | Metal | 36.2 |
| | 13 | Doorbell | Hospital access | Plastic | 36.4 |
| | 14 | Iron grades | Principal entrance | Metal | 38.2 |
| | 24 | Bench | Principal entrance | Wood | 37.3 |
| | 36 | Bench | Hospital access | Wood | 36.3 |
| | 39 | Light switch | Toilet | Plastic | 36.3 |
| | 43 | Wall | Hospital access | Rock | 35.6 |
| | 44 | Wall | Hospital access | Rock | 35.5 |
| | 45 | Handrail | Hospital access | Metal | 37.4 |
| | 46 | Handrail | Hospital access | Metal | 36.6 |
| | 47 | Handrail | Hospital access | Metal | 34.8 |
| | 48 | Handrail | Hospital access | Metal | 36.0 |
| | 49 | Wall | Hospital access | Rock | 36.2 |
| | 51 | Phone | Public phone | Plastic | 38.1 |
| | 52 | Handrail | Hospital access | Rock | 34.8 |
| | 53 | Trafic light button | Traffic light button | Plastic | 34.8 |
| | 54 | Trafic light button | Traffic light button | Plastic | 35.2 |
| | 55 | Trafic light pole | Traffic light button | Metal | 35.3 |
| | 57 | Trafic light button box | Traffic light button | Metal | 36.6 |
| | 59 | Park bench | Bus stop | Concrete | 34.4 |

| | | | | | |
|---|---|---|---|---|---|
| | 60 | Bus stop bench | Bus stop | Metal | 36.7 |
| | 61 | Bus stop ceiling | Bus stop | Plastic | 35.0 |
| | 62 | Trash box | Bus stop | Plastic | 35.0 |
| | 63 | Road sign pole | Resting area | Metal | 35.8 |
| | 67 | Handrail | Resting area | Metal | 37.5 |
| **Transport terminals** | 85 | Faucet | Toilet | Plastic | 34.7 |
| | 86 | Wall | Toilet | Rock | 33.6 |
| | 87 | Bench | Bench | Wood | 37.6 |
| | 88 | Handrail | Handrail | Metal | 36.8 |
| | 89 | Toilet seat | Toilet | Other (Ceramic) | 35.4 |
| | 90 | Column | Bus terminal access | Concrete | 35.9 |
| | 91 | Faucet | Faucet | Metal | 32.2 |
| | 92 | Bench | Bench | Wood | 35.2 |
| | 93 | Handrail | Handrail | Metal | 34.1 |
| | 94 | Faucet | Toilet | Plastic | 34.1 |
| | 95 | Handrail | Handrail | Metal | 36.7 |
| | 97 | Bench | Bench | Plastic | 34.9 |
| | 98 | Handrail | Handrail | Metal | 35.9 |
| | 99 | ATM button | ATM | Metal | 36.0 |
| | 100 | Biometrics sensors | ATM | Plastic | 34.4 |
| | 101 | Box | ATM | Plastic | 35.4 |
| | 102 | Button | ATM | Metal | 33.3 |
| | 103 | Biometrics sensors | ATM | Plastic | 35.0 |
| | 104 | Walls | Bus stop | Metal | 35.4 |
| | 105 | Protection grid | Bus stop | Metal | 36.0 |
| | 106 | Button | Ticket machine | Other (Rubber) | 37.6 |
| | 107 | Screen | Ticket machine | Glass | 33.6 |
| | 108 | Handrail | Handrail | Metal | 34.1 |

| | | | | | |
|---|---|---|---|---|---|
| | 109 | Wall | Toilet | Rock | 33.6 |
| | 110 | Bench | Bench | Concrete | 33.7 |
| | 111 | Roulette | Bus terminal exit | Metal | 33.7 |
| | 112 | Column | Bus terminal access | Concrete | 33.7 |
| | 113 | Wall | Subway station access | Rock | 36.5 |
| | 114 | Handrail | Handrail | Metal | 35.4 |
| | 116 | Biometrics sensors | ATM | Plastic | 36.2 |
| | 117 | Faucet | Faucet | Metal | 36.7 |
| | 120 | Handrail | Handrail | Metal | 36.4 |
| | 122 | Column | Bus terminal access | Concrete | 36.7 |
| | 123 | Pipe | Bus terminal access | Metal | 35.3 |
| | 124 | Protection grid | Bus terminal access | Metal | 36.7 |
| | 125 | Counter | Counter | Rock | 37.6 |
| | 127 | Biometrics sensors | ATM | Plastic | 36.4 |
| | 133 | Bench | Bench | Wood | 37.3 |
| | 134 | Grid | Subway station access | Metal | 35.4 |
| | 138 | Protection grid | Bus terminal access | Metal | 38.7 |
| | 139 | Colunm | Bus terminal access | Concrete | 36.4 |
| | 141 | Handrail | Handrail | Metal | 36.3 |
| | 143 | Screen | Ticket machine | Glass | 36.7 |
| | 144 | Bench | Bench | Wood | 34.6 |
| | 155 | Keyboard | ATM | Metal | 36.4 |
| | 156 | Biometrics sensors | ATM | Plastic | 37.3 |
| | 162 | Handrail | Handrail | Metal | 31.1 |
| **Public parks** | 193 | Toy | Playground | Wood | 37.7 |
| | 197 | Floor | Recreation area | Concrete | 37.1 |
| | 205 | Faucet | Toilet | Metal | 37.3 |
| | 217 | Handrail | Outdoor gym | Metal | 37.3 |

| | | | | | |
|---|---|---|---|---|---|
| | 221 | Bench | Playground | Wood | 37.8 |
| | 229 | Slide | Playground | Metal | 37.3 |
| | 231 | Barbell | Outdoor gym | Metal | 37.2 |
| | 237 | Swing | Playground | Metal | 37.3 |
| | 242 | Wall | Recreation area | Concrete | 36.3 |
| | 249 | Tree trunk | Recreation area | Wood | 36.2 |
| | 281 | Bench | Recreation area | Wood | 37.4 |
| | 288 | Handrail | Handrail | Metal | 37.6 |
| | 305 | Faucet | Toilet | Plastic | 39.7 |
| | 311 | Barbell | Playground | Metal | 37.4 |
| **Public markets** | 319 | Faucet | Toilet | Metal | 37.5 |
| | 320 | Door | Toilet | Wood | 37.9 |
| | 360 | Handrail | Principal entrance | Metal | 36.9 |
| | 386 | Grade | Toilet | Metal | 38.1 |
| **Beach places** | 177 | Tap | Toilet | Metal | 36.1 |
| | 180 | Handrail | Resting area | Wood | 37.0 |
| | 181 | Wall | Toilet | Rock | 37.4 |
| | 188 | Bench | Bench | Rock | 37.9 |
| **Other places** | 258 | Handrail | Handrail | Metal | 38.0 |
| | 261 | Faucet | Toilet | Plastic | 38.7 |

* Samples were numbered 1 to 400. Numbers shown are the ID of the sample tested positive by RT-qPCR.

**Table S4.** Viability of SARS-CoV-2 positive surface samples using the Vero CCL-81 linage.

| ID sample | Material | Swab (Cq value) | P3 – 0h (Cq value) | P3 – 72h (Cq value) | Interpretation |
|---|---|---|---|---|---|
| 86 | Rock | 33.6 | ND | ND | Non-culturable |
| 91 | Metal | 32.2 | ND | ND | Non-culturable |
| 102 | Metal | 33.2 | ND | ND | Non-culturable |
| 107 | Glass | 33.6 | ND | ND | Non-culturable |
| 109 | Rock | 33.6 | ND | ND | Non-culturable |
| 110 | Concrete | 33.7 | ND | ND | Non-culturable |
| 111 | Metal | 33.7 | ND | ND | Non-culturable |
| 112 | Concrete | 33.7 | ND | ND | Non-culturable |
| 162 | Metal | 31.0 | ND | ND | Non-culturable |

P3: Passage 3
